# SARS-CoV-2 variant transmission in a community-health population (Mexico City, Mexico)

**DOI:** 10.1101/2021.10.18.21264783

**Authors:** Wenjuan Zhang, Marcela Martinez, Brian D Davis, Stephanie S Chen, Jorge Sincuir Martinez, Clara Corona, Guadalupe Diaz, Elias Makhoul, Saleh Heneidi, Jorge Goldberg, Jasmine T Plummer, Eric Vail

## Abstract

The SARS-CoV-2 variant, B.1.1.519, arose in North and Central America, circulating primarily in Mexico. We demonstrate that this variant peaked during the second wave of COVID-19 in Mexico City in the spring of 2021. This variant is likely more infectious, attributed to mutation in the RBD of the spike protein T478K also seen in the alpha variant (B.1.1.7). However the time dynamics of the spread of this variant drastically changed upon the introduction of delta (B.1.617.2) to the country in which we observe a shift from 0% in May 2021 to 55% delta in the span of one month. Since the delta variant has dominantly spread across the globe, we investigated the increasing frequency of the Mexico variant, B.1.1.519, in the public community within Mexico City. Once present, the delta variant was 78% of the Mexico City catchment in July 2021, a time which marked the commencement of Mexico’s third wave. Our data supports the growing concern that the delta variant is closely associated with the massive infection spread of the VOC in Central and South America. While the T478K mutation, also seen in the alpha variant, has evidence for increased transmissibility, these data suggest that the delta variant shows overall increased fitness seeing as it outcompeted the B.1.1519 this Mexico community.

## Introduction

Assessing core genomic features of SARS-CoV-2 can be used for comparative analysis as a useful tool to track sources of transmission and SARS-Cov-2 evolution within communities and throughout the world (1). Novel variants of SARS-CoV-2 including those seen in the UK (alpha/20I/501Y.V1/B.1.1.7) (2), South Africa (beta/20H/501Y.V2/B.1.351) (1,3), Brazil (gamma/P.1/20J/501Y.V3/B.1.1.248) (4), India (delta and kappa) (5) and South America (lambda) (6) have emerged with the concern of increased infectivity and virulence (7,8). The rate of evolution of SARS-CoV-2 in 2020 from current estimates is approximately 24-25 substitutions per year (9). With a growing rate of COVID cases, tracking these variants within countries becomes essential for evaluating variants that will rapidly displace other variants and predominate globally.

From previous reports, a variant defined by the T478K variant was detected in Mexico and was reported to be likely more transmissible based on the doubling of cases containing this variant in a short time period from March to April 2021 (10). This variant B.1.1.519 has T478K, D614G, P681H, and T732A in spike protein also seen to be equally represented in both sexes and no clinical observations have been reported.

We sought to track the local community spread within the Mexico City metropolitan area during the height of its second and third wave of COVID-19. By July 2021, Mexico entered its third wave of the SARS-CoV-2 pandemic with infections rising 29% compared to the previous week with over 20,000 new cases daily (11). Historically, Mexico City has been highest, accounting for one third of the country’s positive cases in the first and second wave (12). With low vaccination rates (only 27% of the population fully vaccinated) and >95% of the hospitalized COVID-19 positive patients are unvaccinated (13), understanding and tracking the emerging variants in this vulnerable population. Here we report the fitness of the delta variant and its ability to outcompete other emerging variants, alpha (B.1.1.7) and the Mexico variant, B.1.1.519 in this community.

## Methods

### Diagnostics and Sample preparation

Appropriate regulatory review was completed by the CSMC Office of Research Compliance and Quality Improvement (IRB #629_MOD6887). Samples were collected by nasopharyngeal swabs from patients presenting COVID-19 like symptoms. Total nucleic acid was extracted using the QIAamp Viral RNA Mini Kit on the QIASymphony (Qiagen, Germantown, USA). Following the protocol from our previous study(14), all patients were first assessed by RT-qPCR for SARS-CoV-2 viral RNA for N, E and RdRp gene expression. Samples were diagnostically COVID-19 positive with amplification of the targeted region crossing the threshold before 30 cycles. In total, 480 COVID-19 positive samples were used for parallel NGS analysis.

### Targeted NGS and variant analyses

All samples were processed for NGS and phylogenetic analysis similar to Zhang et al., 2021. All sequencing reads were mapped to SARS-CoV-2 genome and analyzed using our previously established protocol(14,15). Data used in this study has been deposited to GISAID. Phylogenetic analysis with CSMC cohort and a global subsampling was performed. At the time of publication, global SARS-CoV-2 clades were assigned based on pangolin designations and nomenclature based on WHO designations.

### Statistical Analysis

Results are presented by months and age demographics of the cohort. We conducted statistical analyses, including medians and paired t-tests.

## Results

Phylogenetic analysis of a public health community in Mexico city revealed a diverse set of lineages (Figure 1). The largest proportion of individuals carried the B.1.1.519 variant most prominent in Mexico (85%)during the beginning of the second wave (March 2021). While the predominant variant B.1.1.519 was a variant unique to South America/Mexico, the alpha variant was also circulating in this population (8%) at that time. During the second wave, the alpha variant became the predominant variant at 39% in May 2021 and 28% in June 2021. As the third wave commenced in July 2021, the delta variant increased dramatically to 78% of cases and alpha variant was dramatically reduced to almost 0%. This is consistent with what was observed in the larger Mexico country population (81%in July 2021 and 98% by September 2021) (16).

**Figure 1.**
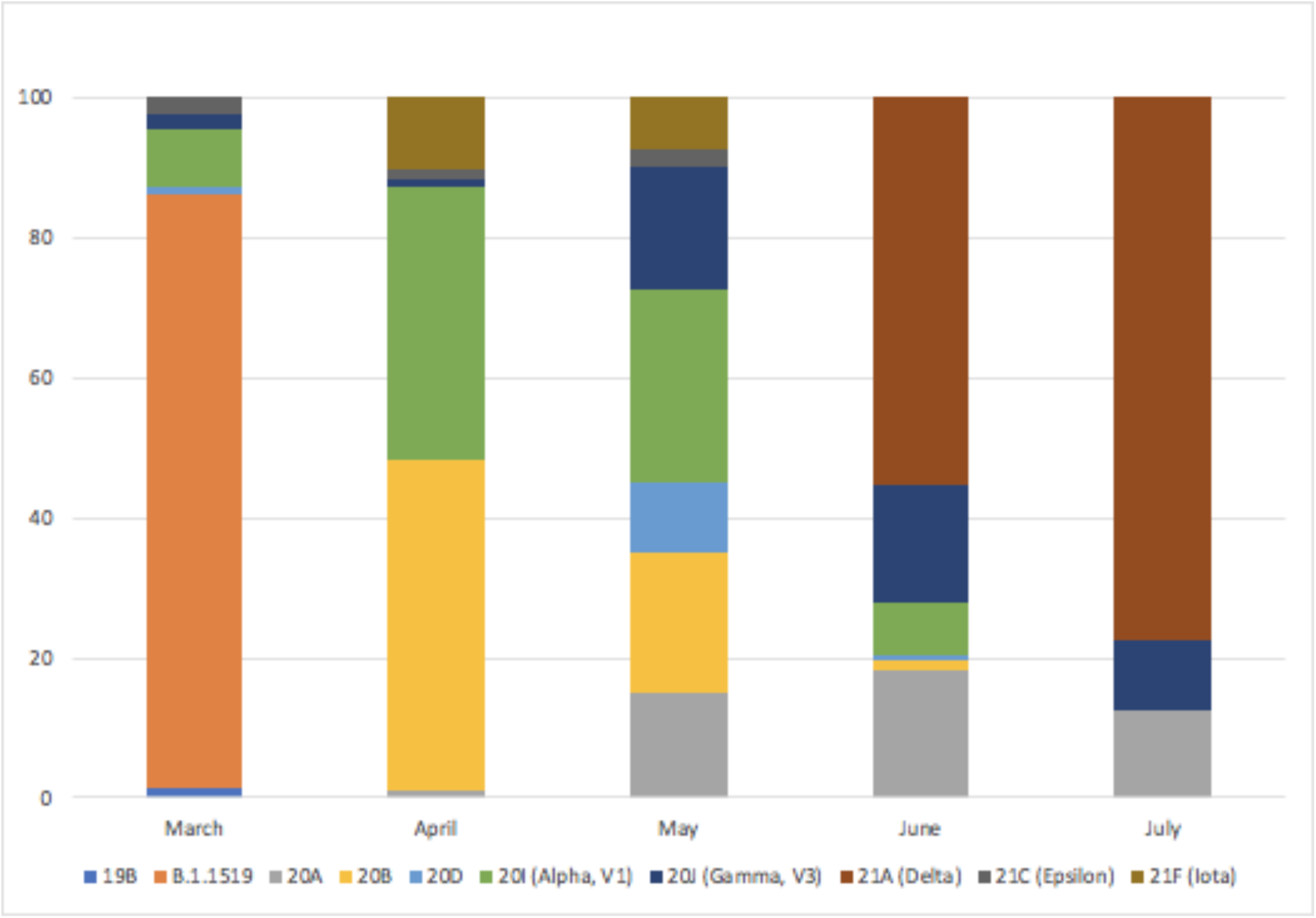
SARS-CoV-2 variant proportions in Mexico city, MX from March 2021 to July 2021.

From all COVID-19 positive samples, we observed younger individuals were affected, with a mean age of 30 years old (Figure 2) and no gender bias was observed in this population (Figure 3).

**Figure 2.**
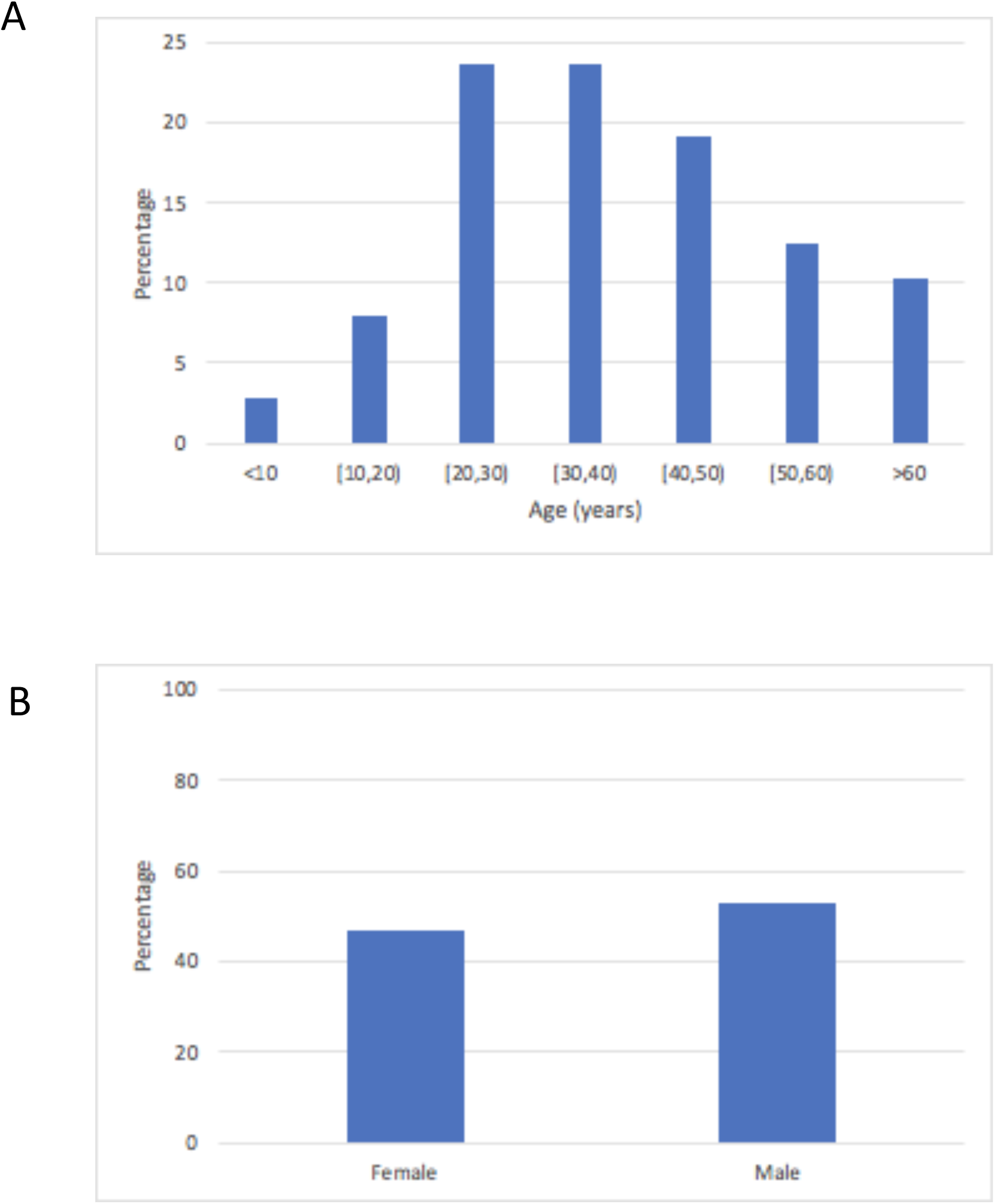
**(A)** Age stratification of COVID-19 positive cases in Mexico City, MX from March 2021 to July 2021. **(B)**. Sex breakdown percentage of COVID-19 positive cases in Mexico City, MX from March 2021 to July 2021.

## Discussion

While alpha and delta variants are both highly transmissible (5,17), our data demonstrates that the delta variant can outcompete the alpha variant in a large metropolitan area. These findings are consistent with what has been reported globally, in which the delta becomes the predominant variant (18) (19) even in populations with a high proportion of COVID-19 individuals carrying the alpha variant. In fact, our data demonstrates the competition of delta versus alpha results in the delta variant being more fit and able to take over both the Mexico and alpha variant within our Mexico City catchment in less than a 4 week time period. We report for the first time that like many emerging SARS-CoV-2, B.1.1.519 was found to consistently affect younger patients. The decrease seen in age of infection is concerning as many young adults and children remain unvaccinated throughout the globe.

The drastic changes of variant proportions from June to July 2021 in this cohort, indicates that though the alpha and B.1.1.519 variant have evolved to be more transmissible, the delta variant has outcompeted other variants in this large metropolitan area within a few weeks. This result highlights the importance of increasing efforts for vaccine rollout in countries who have had limited access to date. With Mexico, having only 26% (12,13)of their country’s population being fully vaccinated, data from this study validate growing concerns that emerging variants such as delta plus and lambda which contain similar mutations to delta, will likely quickly overtake this relatively unprotected population.

## Data Availability

All data produced in the present study are available upon reasonable request to the authors

## Competing Interests

Authors do not have and conflicts of interests to disclose.

## Notes

### Competing Interest Statement

The authors have declared no competing interest.

### Funding Statement

Funding provided by Cedars-Sinai International

### Author Declarations

Appropriate regulatory review was completed by the CSMC Office of Research Compliance and Quality Improvement (IRB #629_MOD6887)

